# Topographical differences in white matter hyperintensity burden and cognition in aging, MCI, and AD

**DOI:** 10.1101/2022.04.20.22274087

**Authors:** Farooq Kamal, Cassandra Morrison, Josefina Maranzano, Yashar Zeighami, Mahsa Dadar, Alzheimer’s Disease Neuroimaging Initiative

## Abstract

**Background:** White matter hyperintensities (WMHs) are pathological changes that develop with increased age and are associated with cognitive decline. Most research on WMHs has neglected to examine regional differences and instead focuses on using a whole-brain approach. This study examined regional WMH differences between normal controls (NCs), people with mild cognitive impairment (MCI), and Alzheimer’s disease (AD). Another goal was to examine whether WMH burden was associated with declines in different cognitive domains in each of the groups.

**Methods:** Participants were selected from the Alzheimer’s Disease Neuroimaging Initiative and included if they had at least one WMH measurement and cognitive scores examining global cognition, executive functioning, and memory. MCI and AD participants were included only if they were amyloid positive. A total of 1573 participants with 7381 follow-ups met inclusion criteria. Linear mixed-effects models were completed to examine group differences in WMH burden and the association between WMH burden and cognition in aging, MCI, and AD.

**Results:** People with MCI and AD had increased total and regional WMH burden compared to cognitively healthy older adults. An association between WMH and cognition was observed for global cognition, executive functioning, and memory in NCs in all regions of interest. A steeper decline (stronger association between WMH and cognition) was observed in MCI compared to NCs for all cognitive domains in all regions. A steeper decline was observed in AD compared to NCs for global cognition in only the temporal region.

**Conclusion:** These results suggest WMH burden increases from aging to AD. A strong association is observed between all cognitive domains of interest and WMH burden in healthy aging and MCI, while those with AD only had a few associations between WMH and memory and WMH and global cognition. These findings suggest that WMH burden is associated with changes in cognition in healthy aging and early cognitive decline, but other biological changes may have a stronger impact on cognition with AD.

## 1 Introduction

Increased age is associated with changes in brain structure and function, which may be typical (i.e., normal) or pathological (i.e., abnormal). These changes may result in the development of cognitive decline, including mild cognitive impairment (MCI) or dementia. Declines in cognitive functioning characterize MCI that are not severe enough to interfere with everyday functioning [1]. When the cognitive decline progress to interfering with everyday functioning, the impairment is then considered dementia [1]. MCI and dementia prevalence increases with aging and are associated with neurodegenerative brain changes [2,3]. Alzheimer’s Disease (AD) is the most common form of dementia, accounting for between 60-80% of dementia cases [3].

One common pathological brain change seen in the aging population is the increase in white matter hyperintensity (WMH) burden [4–6]. WMHs are measured as a proxy of cerebrovascular disease and are observed as areas of increased signal in T2-weighted or fluid-attenuated inversion recovery (FLAIR) magnetic resonance images (MRIs) [7]. WMHs are observed in clinically healthy older adults [4–6]; however, WMHs may be one of the first pathological changes associated with cognitive decline [8]. High WMH burden is an indicator of future cognitive decline [9] and progression to MCI or dementia [6,10,11].

WMH burden is generally increased in each progressive stage of cognitive decline. That is, WMHs are lower in cognitively unimpaired older adults (NC) and progressively increase from MCI to AD [6]. Research examining WMH burden tends to measure global WMH burden as opposed to regional WMH burden (e.g., Brugulat-Serrat et al., 2020; Dadar, Camicioli, et al., 2020; Garnier-Crussard et al., 2022). The studies that have observed healthy older adults and MCI/AD group differences in regional WMH burden have found differences in all regions. For instance, some studies report that the largest NC-AD group difference in WMH volume is observed in the parietal, temporal, or occipital regions [13–15]. However, previous research notes that WMHs are mostly distributed in both the parietal and frontal lobes [15,16], with MCI and AD patients also showing increased WMHs in frontal regions relative to controls [15]. The regional distribution of WMHs has also been suggested to correlate with different etiologies, with anterior WMHs linked to physiological aging and posterior WMHs associated with AD [15,17]. Finally, increases in WMH volumes in both aging and AD have shown associations with future cognitive decline [6,10,11].

Studies examining the relationship between WMH and cognition in aging and AD have shown varying results. Many studies have noted an association between declines in executive functioning [18,19], memory [18,20,21], and global cognition [13,22] with WMH volumes in aging and MCI/AD. While, other studies have observed no association between executive functioning [13], memory [13,23,24], and global cognition [25] with WMHs in aging and MCI/AD. A recent meta-analysis examined 22 studies examining at WMH and cognition in MCI and AD [26]. These authors found a small to medium effect size relationship between cognition and WMH. More specifically, the strongest associations between WMH and cognition were observed with attention, executive functioning, and processing speed. These associations were medium sized in MCI but small in AD. However, the problem with many of the studies in this review is their cross-sectional nature (only 1 was longitudinal). WMHs are known to be more strongly associated with future cognitive decline than immediate cognitive functioning [9]. Therefore, not examining future cognitive decline is a limitation of these studies. Few of the abovementioned studies have assessed regional changes longitudinally and compared these regional changes to cognition.

The progressive increase in WMH burden from healthy older adults to people with MCI and people with AD is relatively well established in research. However, spatial WMH distributions and regional effects on domain-specific cognition remain unknown between the groups. Also unexplored, is how the association between cognitive change over time and WMH burden in MCI and AD differs from that of cognitively normal healthy older adults. Using data from the Alzheimer’s Disease Neuroimaging Initiative (ADNI), the present study will look to answer these questions by investigating the regional accumulation of WMH burden across amyloid-beta positive (A*β*-positive) adults with AD, A*β*-positive adults with MCI (i.e., those on the AD trajectory), and cognitively healthy older adults. The study will also examine the association between cognition and regional WMH across groups, both at baseline and over time.

## 2 Methods

### 2.1 Alzheimer’s Disease Neuroimaging Initiative

The data used in the preparation of this article were obtained from the Alzheimer’s Disease Neuroimaging Initiative (ADNI) database (adni.loni.usc.edu). The ADNI was launched in 2003 as a public-private partnership, led by Principal Investigator Michael W. Weiner, MD. The primary goal of ADNI has been to test whether serial magnetic resonance imaging (MRI), positron emission tomography (PET), other biological markers, and clinical and neuropsychological assessment can be combined to measure the progression of mild cognitive impairment (MCI) and early Alzheimer’s disease (AD). The study received ethical approval from the review boards of all participating institutions. Written informed consent was obtained from participants or their study partner. Participants were included from all ADNI cohorts (ADNI-1, ADNI-2, ADNI-GO, and ADNI-3).

### 2.2 Participants

Participant inclusion and exclusion criteria are available at www.adni-info.org. All participants were between 55 and 90 years of age at the time of recruitment and exhibited no evidence of depression. Healthy older adult controls (NCs) had no evidence of either memory decline, as measured by the Wechsler Memory Scale, or impaired global cognition as measured by the MiniMental Status Examination (MMSE) or Clinical Dementia Rating (CDR). MCI participants obtained scored between 24 and 30 on the MMSE, 0.5 on the CDR, and exhibited abnormal scores on the Wechsler Memory Scale. AD participants exhibited abnormal memory function on the Wechsler Memory Scale, obtained an MMSE score between 20 and 26, a CDR of 0.5 or 1.0, and had probable AD according to the National Institute of Neurological and Communicative Disorders and Stroke and the Alzheimer’s Disease and Related Disorders Association criteria [27]. To ensure that the MCI and AD participants included in this study were in fact MCI due to AD, and AD (as opposed to another form of dementia), they were only included if they were amyloid positive. This selection criterion ensured that both MCI and AD participants were actually on the AD trajectory. Both PET and CSF values were used to determine amyloid positivity in people with MCI and AD because some participants had only one measurement available. Amyloid positivity was identified based on the following criteria: 1) a standardized uptake value ratio (SUVR) of > 1.11 on AV45 PET [28], 2) an SUVR of >1.2 using Pittsburgh compound-B PET [29], 3) an SUVR of ≥1.08 for Florbetaben (FBB) PET [30], or 4) a cerebrospinal fluid Aß1-42 ≤ 980 pg/ml as per ADNI recommendations.

A total of 977 MCI and 372 AD participants had WMHs measurements with 6524 followups. Of those participants, 564 MCI and 259 AD participants were amyloid positive. The amyloid positive MCI and AD participants were included in this study along with the 750 NCs, for a total of 1573 participants with 7381 follow-up time points. Participants were included from all ADNI cohorts including ADNI-3.

### 2.3 Structural MRI acquisition and processing

All participants were scanned consistently using the standardized acquisition protocols provided by the ADNI. Longitudinal MRI data were downloaded from the ADNI public website. For the detailed MRI acquisition protocol and imaging parameters see http://adni.loni.usc.edu/methods/mri-tool/mri-analysis/. T1w scans for each participant were preprocessed through our standard pipeline including noise reduction [31], intensity inhomogeneity correction [32], and intensity normalization into range [0-100]. The pre-processed images were then linearly (9 parameters: 3 translation, 3 rotation, and 3 scaling) [33] registered to the MNI-ICBM152-2009c average template [34]. The quality of the linear registrations was visually verified by an experienced rater (author M.D.), blinded to diagnostic group. Only seven scans did not pass this quality control step and were discarded.

### 2.4 WMH measurements

A previously validated WMH technique that has been extensively tested for assessment of WMHs in aging was used to obtain WMH measurements. This technique has been previously employed in other multi-center studies [8,9,35] and has also been validated in the ADNI cohort [36]. Automatic segmentation of the WMHs was completed using the T1w contrasts, along with location and intensity features from a library of manually segmented scans (50 ADNI participants independent of the ones studied here) in combination with a random forest classifier to detect the WMHs in new images [8,37]. WMH load was defined as the volume of all voxels identified as WMH in the standard space (in mm^3^) and were normalized for head size. Regional and total WMH volumes were calculated based on Hammers Atlas [37,38]. All WMH volumes were also logtransformed to achieve normal distribution.

### 2.5 Cognitive Scores

Participant cognitive scores were also downloaded from the ADNI website. The Alzheimer’s Disease Assessment Scale-13 (ADAS-13) was included as a measure of global cognitive functioning. Ray’s Auditory Verbal Learning test (RAVLT) was included as a measure of episodic memory, and the Trail Making Test Part B (TMT-B) was included as a measure of executive functioning. While other neuropsychological tests were completed throughout ADNI, these specific tests were included because over 80% of the participants completed these assessments.

### 2.6 Statistical Analysis

#### 2.6.1 Baseline Assessments

Participant demographic information is presented in Table 1. T-tests and chi-square analyses were performed on the demographic information and corrected for multiple comparisons using Bonferroni correction. The following linear regression models were used to investigate group differences in total and regional WMH loads, including age, sex, years of education, and APOE4 status as covariates.

**Table 1:** Demographic and clinical characteristics for NCs, MCI and AD

| Demographic Information | NC<br>n = 750 | MCI<br>n = 564 | AD<br>n = 259 |
| --- | --- | --- | --- |
| Baseline Age | 73.2 ± 6.2 | 73.2 ± 7.2 | 74.2 ± 8.01 |
| Education | 16.4 ± 2.6 | 16.0 ± 2.8<sup>1</sup> | 15.6 ± 2.7<sup>1</sup> |
| APOE ε4+ | 230 (31%) | 349 (62%)<sup>1</sup> | 190 (73%)<sup>1,2</sup> |
| Male sex | 339 (45%) | 332 (59%)<sup>1</sup> | 147 (57%)<sup>1</sup> |
| Baseline ADAS13 | 8.9 ± 4.4 | 17.3 ± 6.7<sup>1</sup> | 30.6 ± 8.3<sup>1,2</sup> |
| Baseline TMT-B | 81.6 ± 41.2 | 121.6 ± 68.6<sup>1</sup> | 198.7 ± 85.7<sup>1,2</sup> |
| Baseline RAVLT – Immediate | 45.3 ± 9.9 | 33.0 ± 9.7<sup>1</sup> | 22.4 ± 7.4<sup>1,2</sup> |
| Baseline RAVLT – Learning | 6.0 ± 2.4 | 4.0 ± 2.6<sup>1</sup> | 1.8 ± 1.8<sup>1,2</sup> |
| Baseline RAVLT – Perc – Forgetting | 35.6 ± 22.1 | 64.35 ± 31.1<sup>1</sup> | 88.96 ± 22.1<sup>1,2</sup> |

Notes: values are expressed as mean ± standard deviation, or number of participants (percentage %). <sup>1</sup> = statistically significant difference from NC. <sup>2</sup> = statistically significant difference from MCI. APOE ε4+ = number and percentage of people with at least one ε4 allele. NC = normal control. MCI = mild cognitive impairment. AD = Alzheimer's disease. ADAS13 = Alzheimer's Disease Assessment Scale – 13. TMT-B = Trail Making Test- B. RAVLT = Ray Auditory Verbal Learning Test. Perc – Forgetting = percent forgetting. For baseline ADAS-13, TMT-B, RAVLT-Perc-Forgetting higher scores represent poor performance. For RAVLT-Immediate and RAVLT-Learning lower scores represent poor performance.

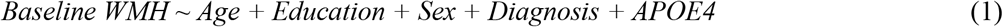

To investigate whether the WMHs were disproportionately distributed across lobes in different diagnostic groups, the lobar WMH volumes (frontal, parietal, temporal, and occipital) were further normalized over the total WMH volume. Similar linear regression models were then completed to investigate whether there were differences in WMH proportions across groups for each lobe.

#### 2.6.2 Clinical-Anatomical Signatures

Partial least squares (PLS) method was used to determine the baseline patterns of association between WMH measures and the cognitive domains in each diagnostic group. PLS is a singular value decomposition based multi-variate method that relates two sets of variables to each other, by finding the linear combinations of the variables that maximally covary with each other [39–41]. We entered WMH measures, as well as age, sex, years of education, and APOE4 status as the contributing factors, and the scores for ADAS-13, TMT-B, and RAVLT learning, immediate and percent forgetting as the cognitive domain variables. 500 bootstraps were performed to calculate the effect sizes and confidence intervals for the loadings of contributing factors and cognitive domains for each latent variable. The p-values for each latent variable were calculated using permutations (N = 500).

#### 2.6.3 Longitudinal Assessments

WMH differences between NC, MCI, and AD were investigated using linear mixed effects models to examine the association between WMH load (frontal, temporal, parietal, occipital, and total) and diagnosis. Regional WMH values (i.e., frontal, temporal, parietal, and occipital) were summed across the right and left hemispheres to obtain one score for each region. All results were corrected for multiple comparisons using false discovery rate (FDR), p-values are reported as raw values with significance then determined by FDR correction [42].

The categorical variable of interest was Diagnosis (i.e., NC, MCI, AD) based on baseline diagnosis, contrasting MCI and AD against the NCs. The model also included age, sex, education, and APOE4 status as covariates. The categorical variable Apolipoprotein E (APOE4) was used to contrast subjects with one or two APOE ε4 alleles against those with zero. Participant ID was included as a categorical random effect.

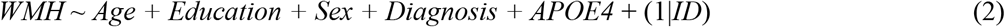

The following linear mixed effects model was conducted to examine whether total or regional WMH would influence cognitive scores. The variables of interest were Diagnosis (contrasting MCI and AD against the NCs), WMH burden, and their interaction (contrasting the slopes of change in cognition in relation to WMH burden for MCI and AD against the NCs). Cognitive Measure represents cognitive scores for global functioning, episodic memory, and executive function.

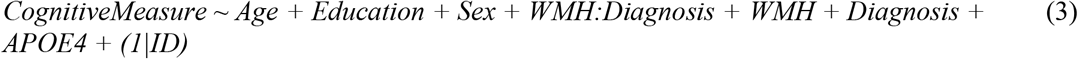

For follow-up measurements, 1578 participants had follow-up scores over a maximum period of 13 years. WMHs were examined in both a regional approach and overall. Analyses were completed separately for WMHs in frontal, temporal, parietal, and occipital and overall. The model also included age, sex, education, and *APOE4 status* as covariates. Participant ID was included as a categorical random effect. All continuous values were z-scored within the population prior to the regression analyses.

All statistical analyses were performed using MATLAB version 2021a.

## 3 Results

### 3.1 Demographics and Cognitive Scores

Table 1 presents demographic information and clinical characteristics of the participants included in the study. NCs had significantly greater education levels than both MCI *(t*=3.32, *p*<.001) and AD *(t*=5.21, *p*<.001), fewer males than both MCI (x^2^= 23.51, *p*<.001) and AD (x^2^=9.84, *p*=.001), and fewer people with APOE ε4+ than both MCI (x^2^= 146.44, *p*<.001) and AD (x^2^=188.62, *p*<.001). There were also fewer APOE ε4+ positive participants in MCI than AD, this effect remained significant after correction for multiple comparisons (x^2^=9.84, *p*=.001).

Baseline cognitive performance decreased with each stage of progressive decline for the ADAS-13 (NC:MCI, *t=-*25.84, *p*<.001; MCI:AD, *t*= -22.41, *p*<.001), TMT-B (NC:MCI, *t=* - 12.23, *p*<.001; MCI:AD, *=* -12.05, *p*<.001), RAVLT learning (NC:MCI, *t=-*14.32, *p*<.001; MCI:AD, *t*= 13.59, *p*<.001), RAVLT immediate (NC:MCI, *t=-*22.40, *p*<.001; MCI:AD, *t*= -17.26, *p*<.001) and RAVLT percent forgetting (NC:MCI, *t=-*17.49, *p*<.001; MCI:AD, *t*= -12.95, *p*<.001)

### 3.2 Baseline Assessments

Figure 1 shows boxplots of baseline WMHs overall and separately for each lobe. As expected, MCI and AD groups had significantly greater WMH burden than the NC group overall and across all regions (*p*<.01, Figure 1, first row). The normalized WMH loads were not significantly different between diagnostic groups across frontal and occipital lobes, whereas parietal WMHs had a disproportionately higher WMH burden (*t*=2.79, *p*=.005) and the temporal WMHs had a disproportionately lower WMH burden (*t*=-3.61, *p*<.001) in the AD group in contrast to NC and MCI (Figure 1, second row).

**Figure 1.**
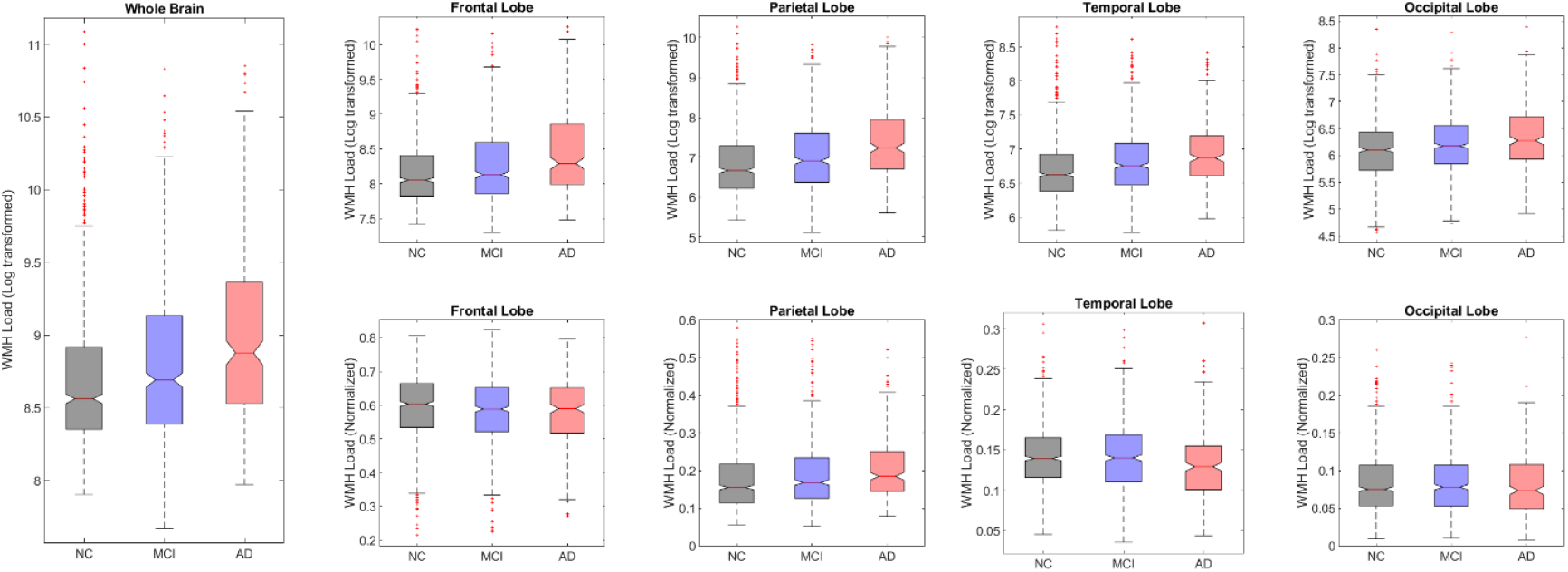
Boxplots showing baseline WMH distributions (log transformed and normalized) across diagnostic groups for each lobe. The first row shows the log transformed WMH loads for each group by lobe. The second row shows the normalized WMH loads for each group by lobe. NC = normal controls. MCI = Mild cognitive impairment. AD = Alzheimer’s Disease. WMH = white matter hyperintensity

#### 3.2.1. Clinical-Anatomical Signatures

The first latent variables were statistically significant for all diagnostic groups (permuted *p*<.05) and accounted for 93.78% of the shared covariance between WMHs and cognitive measures for NC, 85.18% of the shared covariance for MCI, and 52.61 % of the shared covariance for AD. Figure 2 shows the loadings of the contributing factors (left column) and cognitive domains (right column) for the first latent variables in each diagnostic group (rows). In the NCs, greater age, male sex, lower education, and greater WMH burden in all regions were associated with poorer performance in all 5 cognitive domains (indicated by higher scores in ADAS13, TMT-B, and RAVLT percent forgetting, and lower scores in RAVLT immediate and learning). In MCI, a similar pattern emerged, however, RAVLT percent forgetting was no longer statistically significant. In AD, only parietal WMHs, lower education, and RAVLT immediate significantly contributed to the first latent variable.

**Figure 2.**
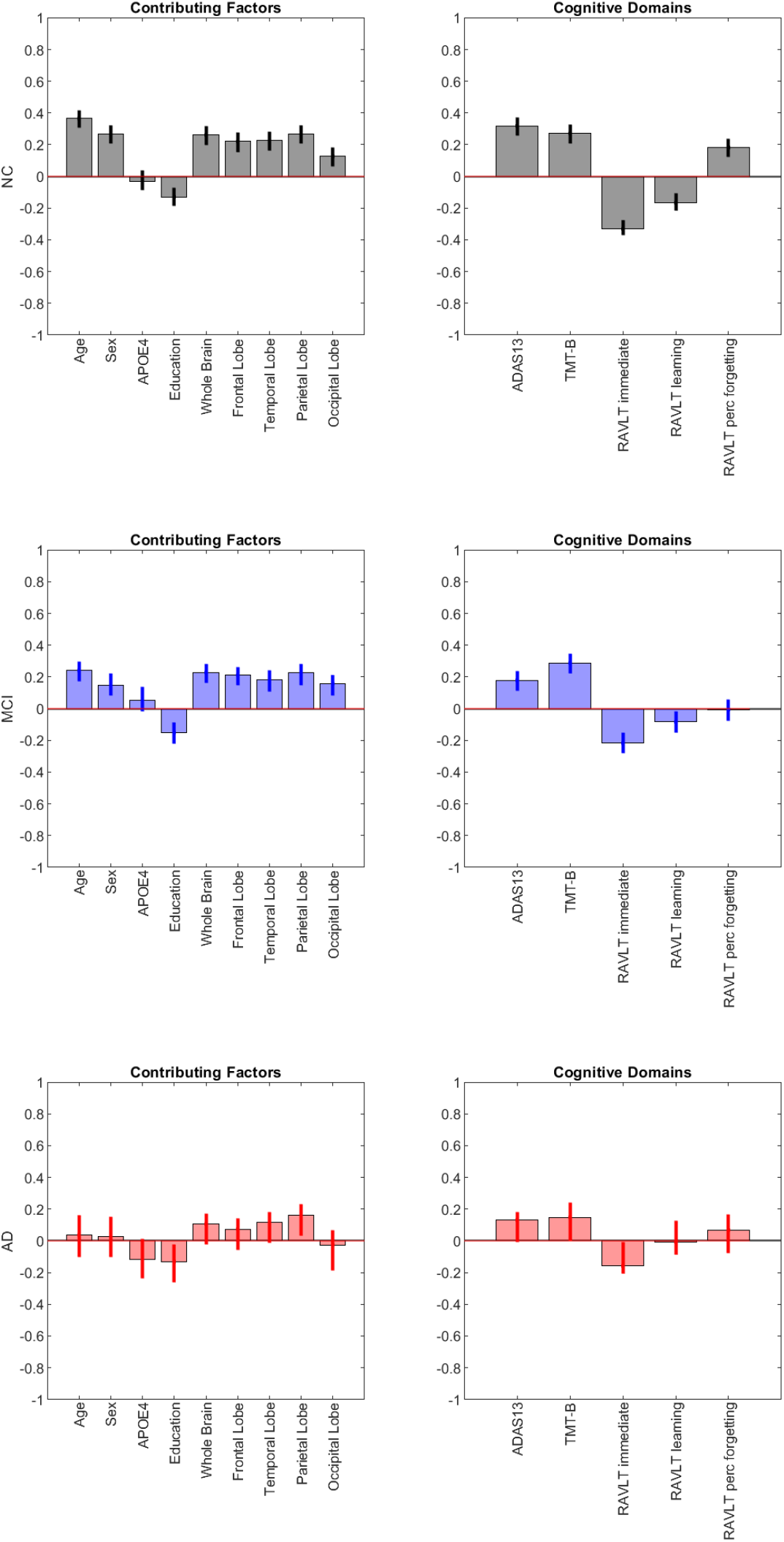
First latent variables from the parietal least squares analysis. Each row represents each diagnosis group, Row 1 = Normal controls, Row 2 = MCI, and Row 3 = AD. The first column shows the contributing factors loadings, while the second column shows the cognitive domains for the first latent variables. NC = normal controls. MCI = mild cognitive impairment. AD = Alzheimer’s disease.

### 3.3 Longitudinal Assessments

#### 3.3.1 WMH analysis of group differences

Table 2 summarizes the results of the longitudinal linear mixed effects models for MCI and AD, contrasted against normal controls across all WMH regions. MCI (*t* belongs to [3.25–5.57], *p*<.001) and AD (*t* belongs to [4.76–7.66], *p*<.001) had significantly increased WMH load in all regions compared to normal controls. At all regions except temporal, people with AD had increased WMH burden compared to MCI (*t* belongs to [3.60–12.18], *p*<.001)

**Table 2:** Linear regression model results showing differences between NCs, MCI, and AD

|  | Total |  | Frontal |  | Temporal |  | Parietal |  | Occipital |  |
| --- | --- | --- | --- | --- | --- | --- | --- | --- | --- | --- |
|  | *T* stat | *P* | *T* stat | *P* | *T* stat | *P* | *T* stat | *P* | *T* stat | *P* |
| **WMH** |  |  |  |  |  |  |  |  |  |  |
| NC vs MCI | 5.50 | **<.001\*** | 5.57 | **<.001\*** | 4.93 | **<.001\*** | 5.07 | **<.001\*** | 3.25 | **.001\*** |
| NC vs AD | 7.29 | **<.001\*** | 7.66 | **<.001\*** | 5.61 | **<.001\*** | 6.60 | **<.001\*** | 4.76 | **<.001\*** |
| MCI vs AD | 10.07 | **.001\*** | 12.18 | **<.001\*** | 3.60 | .057 | 7.85 | **.005\*** | 5.48 | **.019\*** |

Notes: \* Represents results that are significant when uncorrected and bolded values represent those that remained significant after correction for multiple comparisons.

#### 3.3.2 Cognition and regional WMH

Table 3 and Figure 1 summarize the results of the linear mixed effects models examining the longitudinal relationship between cognitive scores and WMH load. For global cognition as measured by ADAS13, there was a significant association with WMH in NC for all regions except temporal (*t* belongs to [3.64–5.33], *p*<.001), demonstrating that increased WMH is associated with increased ADAS13 scores (i.e., worse cognition) in NCs. The interaction between WMH and global cognition was significant for MCI at all regions (*t* belongs to [10.45–12.55], *p*<.001), whereas the interaction between WMH and AD was marginally significant only in the parietal region (*t* =2.18, *p*=.03).

**Table 3:**
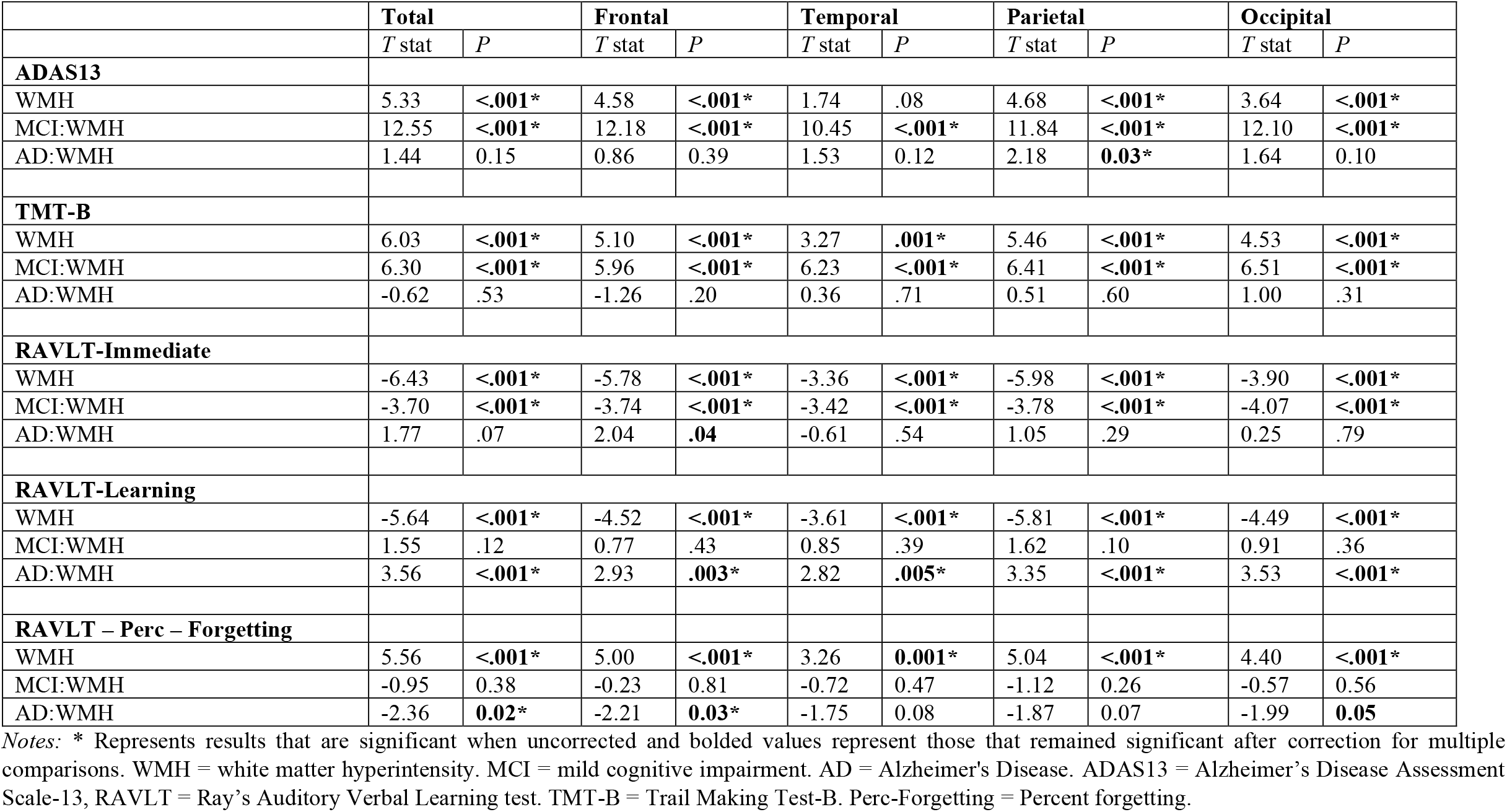
Linear regression model results showing interactions between cognition and WMH at all regions of interest for NCs, MCI, and AD

|  | **Total** |  | **Frontal** |  | **Temporal** |  | **Parietal** |  | **Occipital** |  |
| --- | --- | --- | --- | --- | --- | --- | --- | --- | --- | --- |
|  | *T* stat | *P* | *T* stat | *P* | *T* stat | *P* | *T* stat | *P* | *T* stat | *P* |
| **ADAS13** |  |  |  |  |  |  |  |  |  |  |
| WMH | 5.33 | <.001* | 4.58 | <.001* | 1.74 | .08 | 4.68 | <.001* | 3.64 | <.001* |
| MCI:WMH | 12.55 | <.001* | 12.18 | <.001* | 10.45 | <.001* | 11.84 | <.001* | 12.10 | <.001* |
| AD:WMH | 1.44 | 0.15 | 0.86 | 0.39 | 1.53 | 0.12 | 2.18 | **0.03\*** | 1.64 | 0.10 |
| **TMT-B** |  |  |  |  |  |  |  |  |  |  |
| WMH | 6.03 | <.001* | 5.10 | <.001* | 3.27 | .001* | 5.46 | <.001* | 4.53 | <.001* |
| MCI:WMH | 6.30 | <.001* | 5.96 | <.001* | 6.23 | <.001* | 6.41 | <.001* | 6.51 | <.001* |
| AD:WMH | -0.62 | .53 | -1.26 | .20 | 0.36 | .71 | 0.51 | .60 | 1.00 | .31 |
| **RAVLT-Immediate** |  |  |  |  |  |  |  |  |  |  |
| WMH | -6.43 | <.001* | -5.78 | <.001* | -3.36 | <.001* | -5.98 | <.001* | -3.90 | <.001* |
| MCI:WMH | -3.70 | <.001* | -3.74 | <.001* | -3.42 | <.001* | -3.78 | <.001* | -4.07 | <.001* |
| AD:WMH | 1.77 | .07 | 2.04 | **.04** | -0.61 | .54 | 1.05 | .29 | 0.25 | .79 |
| **RAVLT-Learning** |  |  |  |  |  |  |  |  |  |  |
| WMH | -5.64 | <.001* | -4.52 | <.001* | -3.61 | <.001* | -5.81 | <.001* | -4.49 | <.001* |
| MCI:WMH | 1.55 | .12 | 0.77 | .43 | 0.85 | .39 | 1.62 | .10 | 0.91 | .36 |
| AD:WMH | 3.56 | <.001* | 2.93 | **.003\*** | 2.82 | **.005\*** | 3.35 | <.001* | 3.53 | <.001* |
| **RAVLT – Perc – Forgetting** |  |  |  |  |  |  |  |  |  |  |
| WMH | 5.56 | <.001* | 5.00 | <.001* | 3.26 | **0.001\*** | 5.04 | <.001* | 4.40 | <.001* |
| MCI:WMH | -0.95 | 0.38 | -0.23 | 0.81 | -0.72 | 0.47 | -1.12 | 0.26 | -0.57 | 0.56 |
| AD:WMH | -2.36 | **0.02\*** | -2.21 | **0.03\*** | -1.75 | 0.08 | -1.87 | 0.07 | -1.99 | **0.05** |

Notes: \* Represents results that are significant when uncorrected and bolded values represent those that remained significant after correction for multiple comparisons. WMH = white matter hyperintensity. MCI = mild cognitive impairment. AD = Alzheimer's Disease. ADAS13 = Alzheimer's Disease Assessment Scale-13, RAVLT = Ray's Auditory Verbal Learning test. TMT-B = Trail Making Test-B. Perc-Forgetting = Percent forgetting.

For executive functioning as measured by TMT-B, there was a significant association with WMH in NC for all regions (*t* belongs to [3.27–6.03], *p*<.001; i.e., decreases in executive functioning was associated with increases in WMH burden in all regions). When examining the WMH by diagnosis interaction for executive functioning, there was a significant interaction between MCI and WMH load in all regions (*t* belongs to [5.96–6.51], *p*<.001), whereas the interactions with AD were not significant in any region (*p*>.05). That is, the slopes of changes in executive functioning score associated with WMHs were significantly steeper in MCI (for all regions) but not AD compared to NCs (Figure 3, second row).

**Figure 3.**
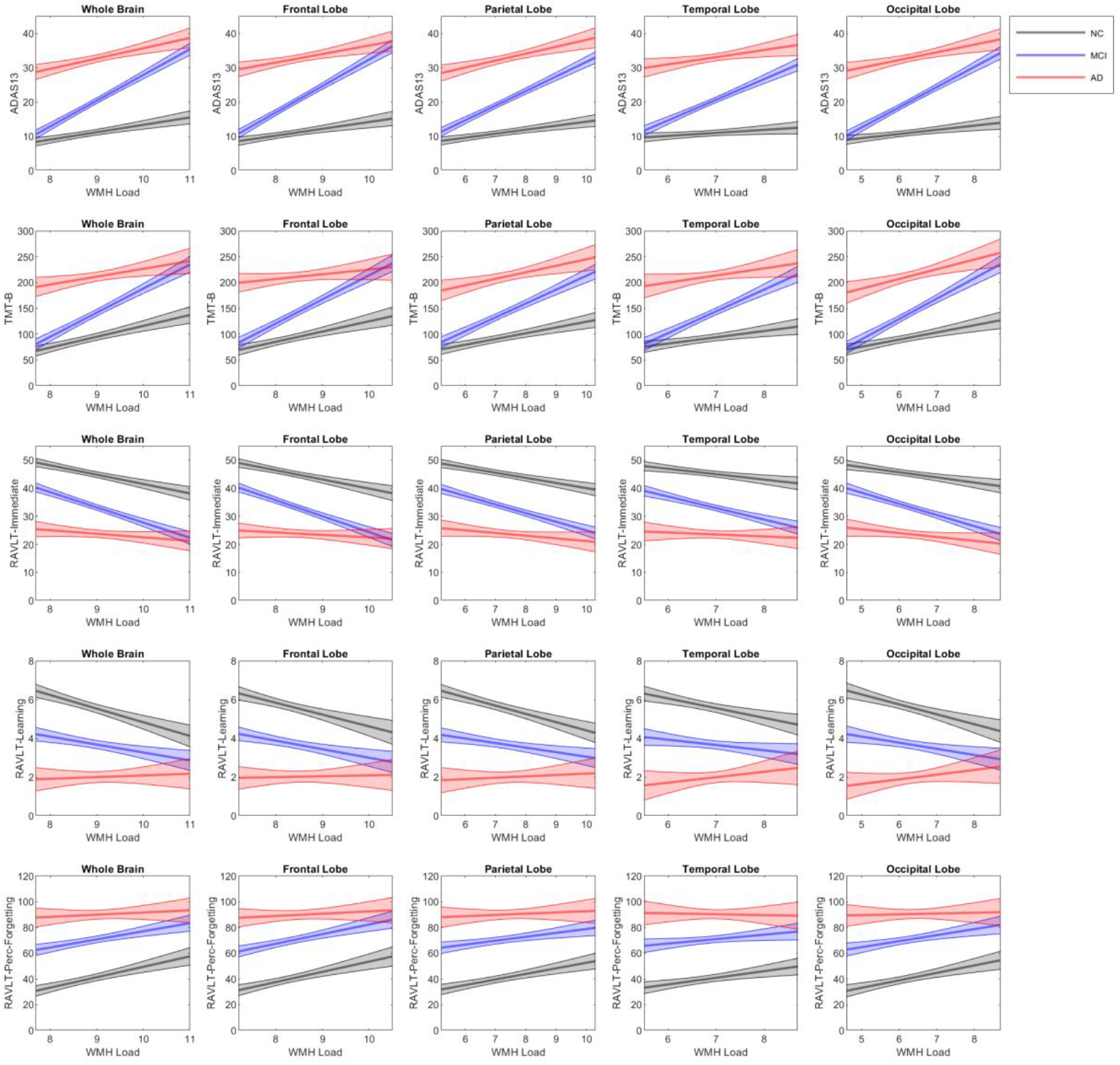
Figures showing the interactions between cognition and WMH at all regions of interest for NCs, MCI, and AD. *Notes:* WMH = white matter hyperintensity. MCI = mild cognitive impairment. AD = Alzheimer’s Disease. ADAS13 = Alzheimer’s Disease Assessment Scale-3, RAVLT = Ray’s Auditory Verbal Learning test. TMT-B = Trail Making Test-B. Perc-Forgetting = Percent forgetting.

For memory as measured by RAVLT (immediate, learning, and percent forgetting), there was a significant association with WMH in NC for all regions (*t* belongs to [3.26–6.43], *p*<.001). For NC, increased WMH load was associated with poorer memory performance in all memory measures. When examining the WMH by diagnosis interaction for RAVLT, there was a significant interaction for MCI and all WMH regions for RAVLT immediate (*t* belongs to [-3.42– -4.07], *p*<.001), but not learning and percent forgetting. For MCI, the slopes of changes in RAVLT immediate were associated with changes in WMH that were significantly steeper than NCs in all regions (Figure 3, third row). AD interaction with WMH burden was not significant in any region (*p*>.05) for RAVLT immediate. AD significantly interacted with WMH across all regions (*t* belongs to [2.82–3.56], *p*<.005) for RAVLT learning, although in the opposite direction, indicating that while WMH burden significantly impacted change in RAVLT learning scores for the NCs, this was not the case in the AD group (note the differences in the slopes in Figure 3, row 4). AD significantly interacted with WMH in the frontal region (*t=*-2.21, *p*=.03) and total (*t*=-2.36, *p* = 0.02) for RAVLT percent forgetting, but again in the opposite direction, indicating a significantly less steep slope in the AD group compared with the NCs. The less steep slopes for AD in comparison with the NC is likely due to the fact that the AD group has reached ceiling on poor performance, as reflected in the much higher intercept values for ADAS13, TMT-B, and RAVLT percent forgetting (for which a higher score indicates poorer performance), and lower values for RAVLT immediate and learning (for which a lower score indicates poorer performance).

## 4 Discussion

The relationship between WMH and future cognitive decline in aging and cognitive impairment is well established. However, the association between cognitive changes and WMH in specific regions is relatively unexplored and inconsistent. Furthermore, whether cognitive changes are influenced differently by WMHs in aging vs. MCI/AD is relatively unknown. The current study was designed to 1) investigate the regional accumulation of WMH burden in healthy older adults, adults with MCI, and adults with AD, and 2) examine the relationship between change in WMH burden and change in cognitive functioning in each group. The study also investigated the impact of regional WMH on cognition across groups. The findings from the present study show that MCI and AD had significantly more WMH burden in all regions compared to NCs. In contrast to NCs, smaller changes in WMH burden in all regions results in higher impact on declines in global functioning, executive functioning, and RAVLT immediate scores in MCI, as reflected by the steeper slopes observed in MCI.

Compared with the NCs, MCI and AD had increased WMH load in all regions as well as in the whole brain analysis. Research has similarly shown an increase in WMH burden at each progressive stage of cognitive decline [6,10,11]. These findings suggest that an increase in widespread WMH pathology occurs in MCI and AD. When examining group WMH differences regionally, studies have typically observed increased WMH in MCI and AD compared to NC in all regions [6,10,11]. In line with previous reports, we found a disproportionately greater prevalence of normalized WMHs in the parietal lobe for the AD group. A larger parietal difference is to be expected because parietal WMHs are reflective of AD pathology as opposed to frontal WMHs which are more reflective of normal aging changes [15,17,44].

Several previous studies have observed an association between cognitive functioning and WMH burden in aging, cognitive decline, MCI, and AD. However, the results of these studies have been inconsistent and most examine total instead of regional WMH burden [13,18,20– 22,24,25]. Our results show that change in cognition (global functioning, executive functioning, and memory) for NCs was associated with total, frontal, temporal, parietal, and occipital WMH burden. People with MCI had an increased change in cognition, which was associated with WMHs, that was above what was observed in NC for all cognitive domains, except two components of memory (i.e., RAVLT learning and RAVLT percent forgetting). That is, MCI had increased slopes compared to NC, for all regions and all domains except RAVLT learning and percent forgetting. When examining people with AD, there were fewer interactions between cognition and WMHs. Compared to NCs, AD only had steeper slopes (i.e., increased change in cognition associated with WMHs) for ADAS-13 and parietal region. For RAVLT percent forgetting and learning, AD exhibited a less steep slope than NCs (Figure 1). The less steep slope observed in AD compared to NCs was not unexpected, because the trajectories inevitably flatten out when the performance of all the participants in the AD group falls in the saturation range of the cognitive scores (see the estimated trajectories for RAVLT percent forgetting as an example -last row in Figure 2- for which the AD group started from an average baseline score of 88.96 -Table 1-, without much further range remaining to detect worsening in performance). That is, the AD participants perform so poorly that the ceiling is reached at baseline, therefore there is less change in scores over time. This interpretation could explain why other studies have observed minimal associations in AD between cognition and WMHs [13,23,24]. These findings suggest WMHs are associated with increased cognitive decline in early aging and prodromal AD (MCI), but only slightly related to changes in AD as detectable by the cognitive batteries currently used. Another potential interpretation that would explain the differences in slopes across the three groups pertains to the presence and impact of other pathologies (i.e., amyloid, tau, and hippocampal neurodegeneration) which may be more strongly related with cognitive deterioration in participants with AD than WMHs. In other words, increased WMH burden contributes to early cognitive decline in aging, but the acceleration in cognitive deterioration, associated with smaller volumetric increases in WMHs, experienced by AD participants, could be driven by other pathological changes in the brains, which are superimposed to the WMHs. To examine the influence of hippocampal atrophy, we repeated these analyses adding hippocampal volume as a covariate. The current results obtained did not change to reflect the influence of neurodegeneration, therefore, these results here likely reflect ceiling effects in AD performance.

Future research should further explore the limitations of this study. Participants in the ADNI dataset have relatively high education levels, and thus may have high cognitive reserve compared to other populations. Future research should aim to examine the influence of WMHs in a population with lower education to determine whether high education levels are protective against cognitive decline caused by WMH burden. The AD participants were in the saturation range of the cognitive scores and therefore the memory test employed here (RAVLT) was not able to capture further worsening of this groups’ cognition. Future research should employ other cognitive batteries that can better capture cognitive change in those with AD. Several other cognitive tests and domains should be examined to better understand the associations between WMH and cognitive change. For example, the Montreal Cognitive Assessment (MoCA) which is sensitive to cognitive changes in MCI and AD [45] may provide additional insight into the relationship between global cognition and WMHs. However, over 50% of our sample was missing MoCA scores and thus MoCA could not be examined in this study.

In addition to these limitations, there are a few strengths of this study that should also be noted. The image processing tools used to analyze the data in this study were previously designed and validated for use in multi-center and multi-scanner studies. These methods have been extensively used to examine neurodegeneration changes in aging, MCI, and dementia [9,46]. The sample size used in this study is very large, with an average of 4.7 follow-up timepoints per participant. The large sample size and follow-up periods improve our statistical power and confidence that the results observed here are an accurate representation of the association between WMH changes with cognition. Finally, our study limited the MCI and dementia patients to those who had confirmed amyloid positivity (using either PET or CSF). This approach is important to show that the dementia participants had an AD diagnosis and that those with MCI were on the AD trajectory. This approach of focusing on only those on the AD trajectory may also explain why some of our results do not match other studies.

## Data Availability

All data used in this article are available from the ADNI public database upon written request for the data.

## Acknowledgments

Data collection and sharing for this project was funded by the Alzheimer’s Disease Neuroimaging Initiative (ADNI) (National Institutes of Health Grant U01 AG024904) and DOD ADNI (Department of Defense award number W81XWH-12-2-0012). ADNI is funded by the National Institute on Aging, the National Institute of Biomedical Imaging and Bioengineering, and through generous contributions from the following: AbbVie, Alzheimer’s Association; Alzheimer’s Drug Discovery Foundation; Araclon Biotech; BioClinica, Inc.; Biogen; BristolMyers Squibb Company; CereSpir, Inc.; Cogstate; Eisai Inc.; Elan Pharmaceuticals, Inc.; Eli Lilly and Company; EuroImmun; F. Hoffmann-La Roche Ltd and its affiliated company Genentech, Inc.; Fujirebio; GE Healthcare; IXICO Ltd.; Janssen Alzheimer Immunotherapy Research & Development, LLC.; Johnson & Johnson Pharmaceutical Research & Development LLC.; Lumosity; Lundbeck; Merck & Co., Inc.; Meso Scale Diagnostics, LLC.; NeuroRx Research; Neurotrack Technologies; Novartis Pharmaceuticals Corporation; Pfizer Inc.; Piramal Imaging; Servier; Takeda Pharmaceutical Company; and Transition Therapeutics. The Canadian Institutes of Health Research is providing funds to support ADNI clinical sites in Canada. Private sector contributions are facilitated by the Foundation for the National Institutes of Health (www.fnih.org). The grantee organization is the Northern California Institute for Research and Education, and the study is coordinated by the Alzheimer’s Therapeutic Research Institute at the University of Southern California. ADNI data are disseminated by the Laboratory for Neuro Imaging at the University of Southern California.

## Conclusion

The findings from this study show a strong relationship between WMH burden changes and cognition in aging and MCI, but not in AD. WMHs were associated with global cognition, executive functioning, and memory in aging and MCI. These findings suggest that WMHs may be a sensitive measure of progressive changes in global cognition, executive functioning, and memory in aging and prodromal AD, but not in later stage AD. Future work is needed to determine if WMHs can predict cognitive decline in healthy older adults and conversion to AD.

